# Burnout among hospital doctors working at a West Midlands Major Trauma Centre: a cross-sectional study

**DOI:** 10.1101/2025.01.14.25320313

**Authors:** Alexandro Basso, Amy Attwater

## Abstract

**Introduction:** Worryingly, burnout is becoming increasingly common among the doctor cohort across the United Kingdom (UK) according to the General Medical Council (GMC), which has also seen an increasing desire to leave the UK profession. The main aims of this study were to: 1) determine the prevalence of burnout among doctors; 2) investigate the association between socio-demographic variables, including thoughts on leaving the profession or the NHS, and both burnout and its two dimensions; 3) examine the potentially predictive relationship between socio-demographic variables and burnout.

**Method:** This cross-sectional study employed the Oldenburg Burnout Inventory (OLBI) to determine the prevalence of burnout as measured across its two core dimensions, exhaustion and disengagement, among doctors working at University Hospital Coventry, a major trauma centre located within the West Midlands. In total, 156 doctors took part in a completed anonymised online questionnaire which also gathered socio-demographic information.

**Results:** 111 (71%) participants were categorised as experiencing burnout, 22 (14%) and 7 (4%) were categorised as exhausted or disengaged, and 16 (10%) were categorised as non-burnout. Whilst there were no associations found between burnout group and socio-demographic variables, there were significant associations found between the exhaustion and disengagement scores and professional grade (*p* < 0.05 and *p* < 0.001, respectively). Doctors who were in the earlier stages of their career, particularly resident doctors in lower training, were found to have significantly higher exhaustion and disengagement scores, and this finding was supported by correlation analysis. There was significant association between burnout group and thoughts on leaving the profession or the NHS (*p* < 0.001). The predictive relationship between socio-demographic variables and burnout groups was found to be poor.

**Conclusion:** At University Hospital Coventry, burnout is a persistent and prevalent issue that affects doctors across all societal and demographic groups. Given the current workforce crisis, more targeted interventions need to be done by organisations to minimise burnout across its two core dimensions to ensure current workforce trends are reversed.

## Introduction

Since the introduction of burnout-related questions in 2018 by the General Medical Council (GMC) in their national training survey, risk of burnout has been demonstrated to have consistently increased until it reached its peak in 2023 (General Medical Council, 2024a). Despite the slight decrease in risk of burnout among the cohorts involved in the 2024 national training survey, risk of burnout continues to remain high with nearly two-thirds (65%) of trainee doctors, who are defined as those in a GMC approved training post, reporting they always or often feel worn out at the end of their day and over half (52%) reporting their work is emotionally exhausting to a high or very high degree. Responses to these questions by trainers, who are defined as postgraduate clinical and educational supervisors involved in the supervision, management, or performance of a trainee, were similarly negative, with over half (52%) of all trainers reporting they always or often feel worn out at the end of the working day.

Given these findings and the current workforce crisis the National Health Service (NHS) which is seeing an increasing number of doctors who are now more likely than ever to leave the United Kingdom (UK) medical profession (General Medical Council, 2024b), it is important to consider whether burnout may play a role in a doctor’s decision to leave the UK medical profession.

It is crucial, therefore, that burnout or risk of burnout is appropriately measured and assessed to ensure a more nuanced evaluation among the sample population. While the GMC national training survey does compare risk of burnout between trainers and trainees, including comparisons between secondary care trainers and general practice (GP) trainers, and analyses risk of burnout by specialty and country, there is no comparison between risk of burnout across different age groups, gender identities, or professional grades. Furthermore, the GMC’s national training survey utilises a subset of the Copenhagen Burnout Inventory (CBI) score to determine risk of work burnout. The CBI focuses on burnout in a single dimension, exhaustion, and measures the extent to which exhaustion is a result of three subscales totalling nineteen questions; personal burnout, work burnout, or client-related burnout (Kristensen et al., 2005).

Though these subscales may be used independently of each other, several studies have shown that healthcare professionals have high scores in both the work burnout and client-related burnout domains (Aiello et al., 2022; Cordero-Franco et al., 2022). The GMC’s decision to focus their remit solely on the seven question work burnout subscale is a limitation worth noting and it may be possible they choose to expand the scope of their work in this area in the future to adequately and more holistically capture the risk of burnout among doctors working in the NHS. This study, however, opted to use the widely validated Oldenburg Burnout Inventory (OLBI) (Halbesleben and Demerouti, 2005; Khan and Yusoff, 2016; Tipa, Tudose, and Pucarea, 2019; Nwosu et al., 2020) as the primary tool to measure burnout among doctors for the following reasons. Firstly, the sixteen question OLBI focuses on burnout across its two core dimensions, exhaustion and disengagement, which allows for a more comprehensive and detailed assessment of burnout. Exhaustion, in this tool, is defined as a consequence of intense physical, affective, and cognitive strain, while disengagement refers to distancing oneself from one’s work or work content (Demerouti and Bakker, 2008). Secondly, the CBI includes only positively phrased questions whilst the OLBI includes both positively and negatively phrased questions. This helps reduce the influence of response bias by introducing cognitive ‘speed bumps’ that ensure a respondent engages with the questions in a more controlled manner (Podsakoff et al., 2003)

With this in mind, the specific objectives of this study were to: 1) determine the prevalence of burnout among doctors working at a major West Midlands trauma centre by using the OLBI; 2) investigate the association between socio-demographic variables, including thoughts on leaving the profession or the NHS, and the findings from the OLBI; 3) examine the potentially predictive relationship between socio-demographic variables and burnout.

## Methods

### Design and setting

A cross-sectional study consisting of an online questionnaire of doctors working at University Hospital Coventry, a West Midlands major trauma centre, was conducted.

### Participants

Potential participants were recruited from University Hospital Coventry between 01/07/2024 to 01/10/2024 via their professional email address and posters containing a QR code linking the participant to the online questionnaire. As this is one of the largest training hospitals in the region it was not possible to determine the exact number of doctors contacted as the number fluctuates due to training rotations and temporary staff changes. Any doctor working at this institution was eligible for inclusion in this study. Participation in this study was completely voluntary and opt-in consent was obtained from all participants prior to their inclusion in the study.

The sample size for this study was calculated using G*Power Software, and used a statistical significance level of 0.05, a study power of 0.8, and an effect size of 0.30 (Faul et al., 2009) This recommended a sample size of 150. 156 participants took part in this study.

### Ethical considerations

Ethical approval for the study was obtained from the University of Warwick’s Biomedical & Scientific Research Ethics Committee (BSREC) and the University Hospital of Coventry and Warwickshire (UHCW) Governance Arrangements for Research Ethics Committees (GAfREC).

### Outcome measures

After collecting socio-demographic information on participants’ gender identity, ethnicity, age group, professional grade, and specialty, participants were presented with an online questionnaire that consisted of the OLBI. The OLBI, as seen in Appendix A, evaluates burnout in its two dimensions, exhaustion and disengagement, with each dimension consisting of eight questions with response options ranging from 1 (strongly agree) to 4 (strongly disagree). Half of the questions presented were negatively phrased and their scores reversed to ensure a higher score indicates more burnout. In this study, the Cronbach’s alpha was 0.86 for exhaustion and 0.83 for disengagement, indicative of a high internal consistency in the responses to each set of questions. In addition to the OLBI, a question was also presented which explored to what extent the participant agreed with the following statement: “Have there been thoughts regarding contemplation of leaving the profession or the NHS?”. Response options included “strongly agree”, “agree”, “neither agree nor disagree”, “disagree”, and “strongly disagree”.

### Data analysis

The online platform Qualtrics was used host the questionnaire which was distributed via email and posters containing an invitation link. Analysis was completed using the statistical package for social sciences (SPSS) software version 29 (IBM Corp, 2023). Categorical data is expressed using frequency and percentage, while continuous data is expressed using mean and standard deviation.

To ensure all professional grades for doctors working in their primary role were appropriately captured, the following categories were used: foundation doctors in years 1 and 2 were categorised as “Foundation Doctor”; resident doctors in core or specialty training years 1 to 3 were categorised as “Resident Doctor in Lower Specialty Training”; resident doctors in specialty training years 4 to 8 were categorised as “Resident Doctor in Higher Specialty Training”; doctors primarily employed as locum, specialist, associate specialist, or specialty doctors, or those locally employed, were categorised as “Locally Employed Doctor/Locum/SAS”; and consultants were categorised as “Consultant.” Similarly, to ensure a comprehensive representation of specialty across participants, the following categories were used: all internal medical specialties were categorised as “Medicine”; all surgical specialties were categorised as “Surgery”; emergency medicine and anaesthetics were categorised as “Emergency Medicine” and “Anaesthetics” respectively; and specialties not fitting into the prior categories such as psychiatry, paediatrics, or radiology were categorised as “Other”.

One-way analysis of variance (ANOVA) was applied to assess differences across socio-demographic variables, including thoughts of leaving the profession or the NHS, by using the mean combined exhaustion and disengagement scores.

Participants were categorised into four burnout groups (burnout, exhausted, disengaged, non-burnout) based on the mean exhaustion and disengagement scores. These groups were determined using the cut-off scores of ≥ 2.25 for exhaustion and ≥ 2.10 for disengagement as used in previous, comparable studies (Schaufeli et al., 2001; Peterson et al., 2008). Scores meeting this criterion are considered high. Participants with high scores of both exhaustion and disengagement levels are categorised as being in the ‘burnout’ group, while high scores in either exhaustion or disengagement are categorised as ‘exhausted’ or ‘disengaged’. Low scores in both dimensions are categorised as ‘non-burnout’. Regression analysis was undertaken to investigate the relationship between burnout groups and socio-demographic variables, including thoughts of leaving the profession or the NHS.

## Results

In total there were 156 participants who took part in the study, most participants were male (*n* = 85, 54.8%), of White ethnicity (*n* = 98, 62.8%), and working in a substantive role as a consultant (*n* = 92, 59.4%). Mean exhaustion and disengagement scores ranged from 1 to 4. In this study, the mean exhaustion score for all participants was 2.82 and the mean disengagement score was 2.47 (Table 1), which are both over the cut-off scores. Figure 1 and Figure 2 show the distribution of these responses.

**Table 1.**

| **OLBI Dimension** | ***M*** | ***SD*** |
| --- | --- | --- |
| *Exhaustion* | 2.82 | 0.59 |
| *Disengagement* | 2.47 | 0.57 |

**Figure 1:**
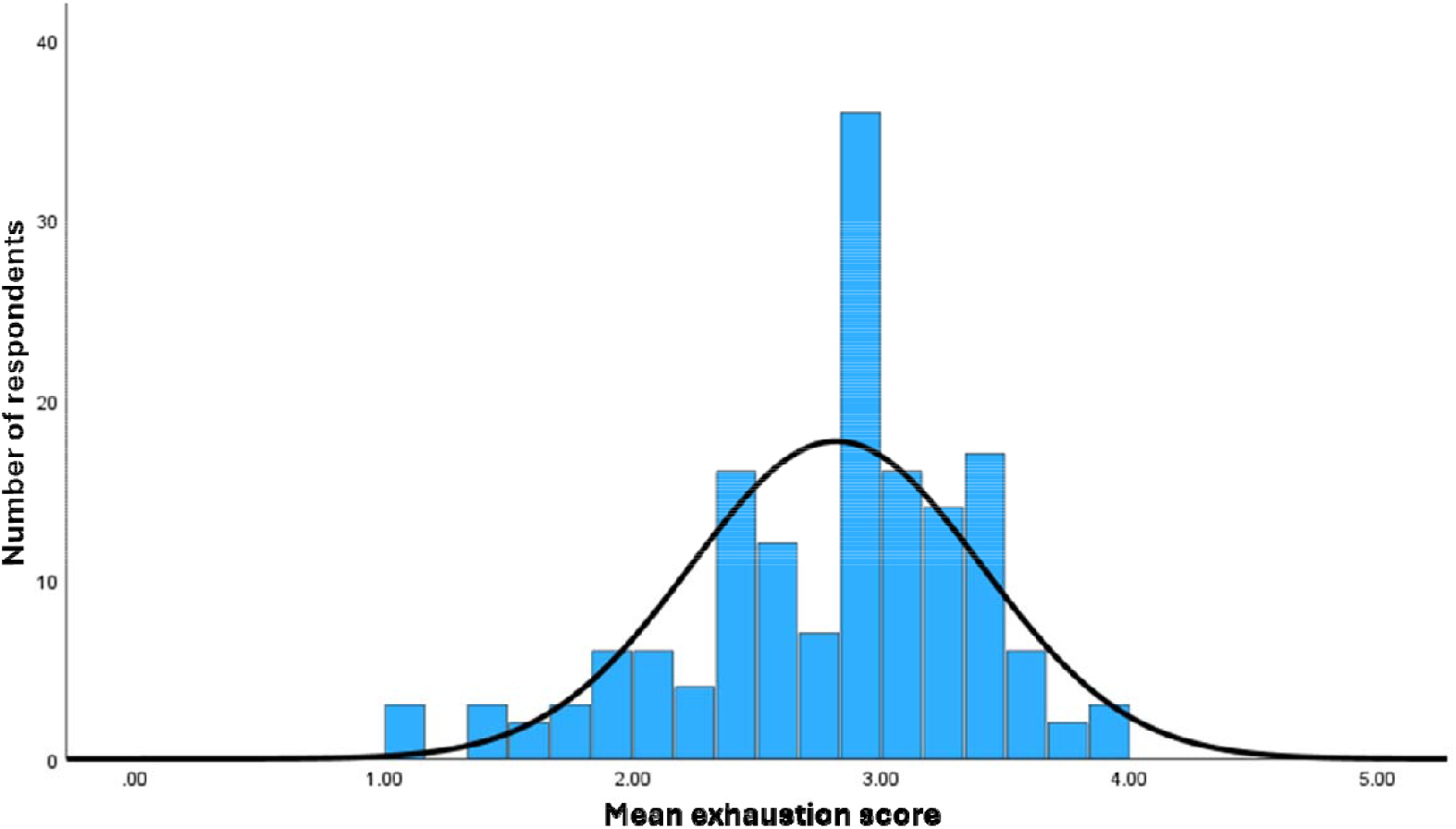
Frequency distribution of mean exhaustion scores.

**Figure 2:**
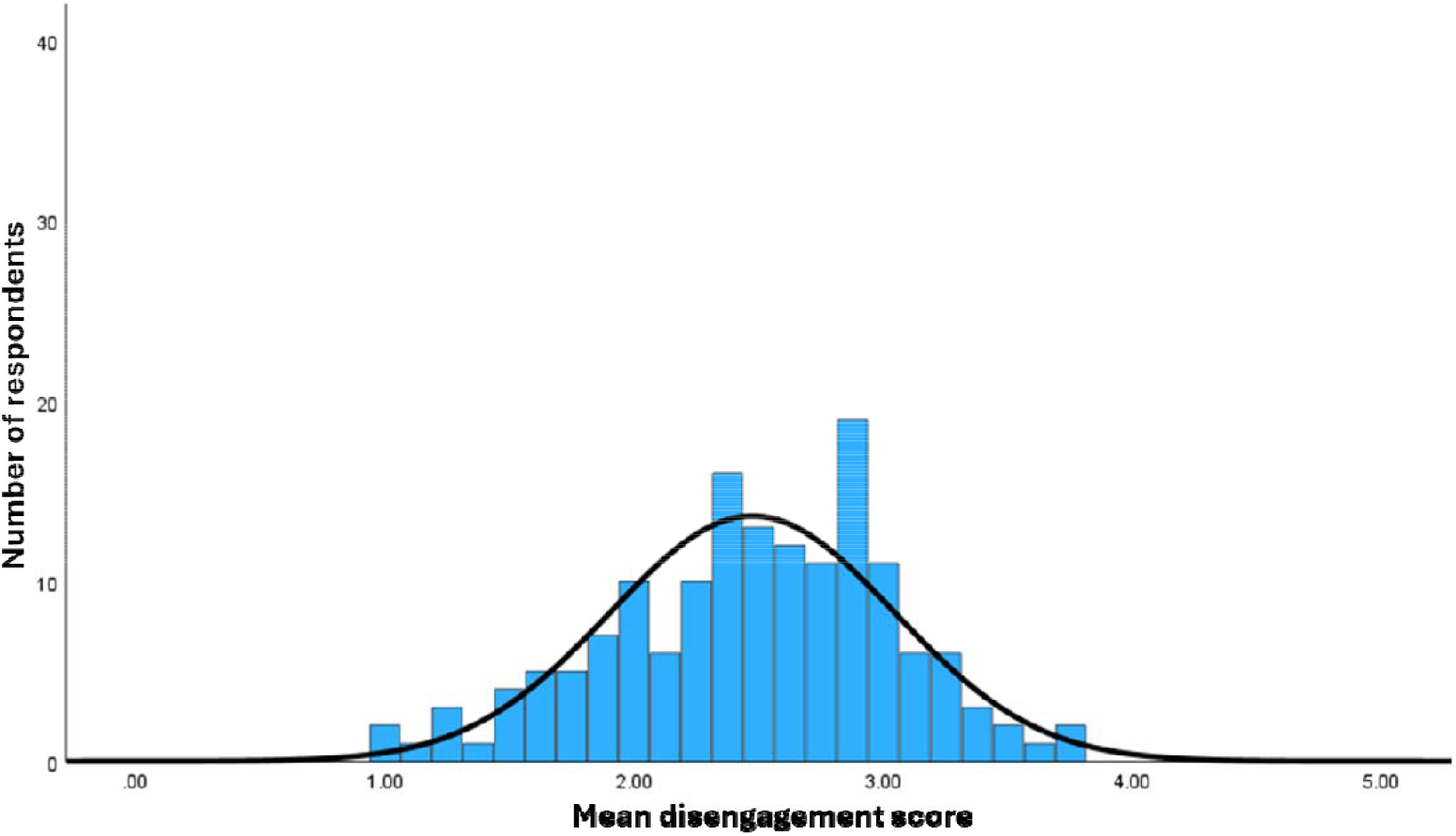
Frequency distribution of mean disengagement scores.

Table 2 shows a full breakdown of the socio-demographic information, including thoughts of leaving the profession or the NHS, and contains the mean exhaustion and disengagement scores, along with one-way ANOVA and, if necessary, post-hoc analysis. Unfortunately, due to an error in the questionnaire allowing participants to continue with the survey without answering all questions, there were two individual responses which did not provide responses to their gender and professional grade. Little’s test of missingness was calculated for these variables and this showed p-values of greater than 0.05 indicating the missing data was missing completely at random. To account for these missing values pair-wise deletion was used during the analysis.

**Table 2.**
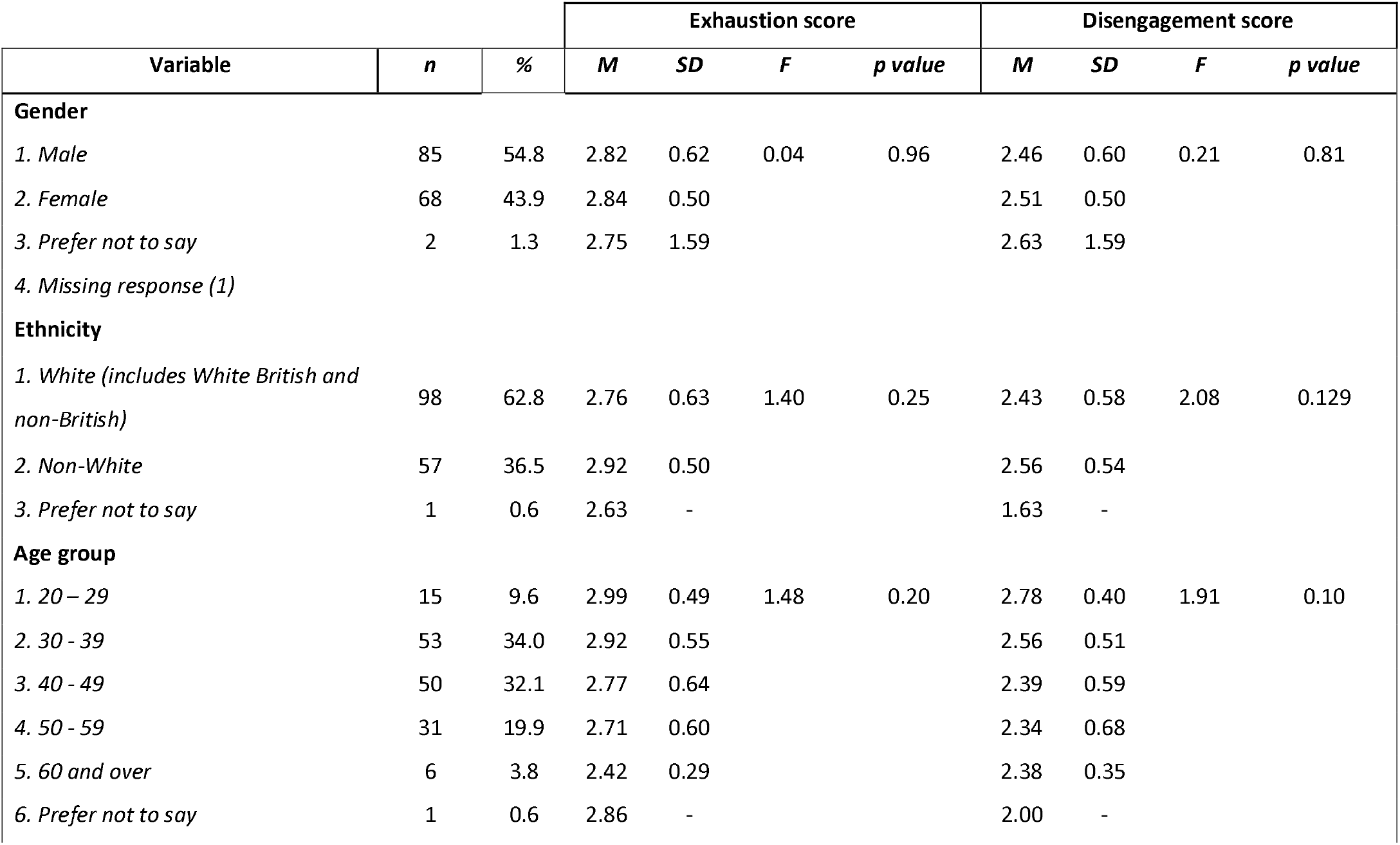
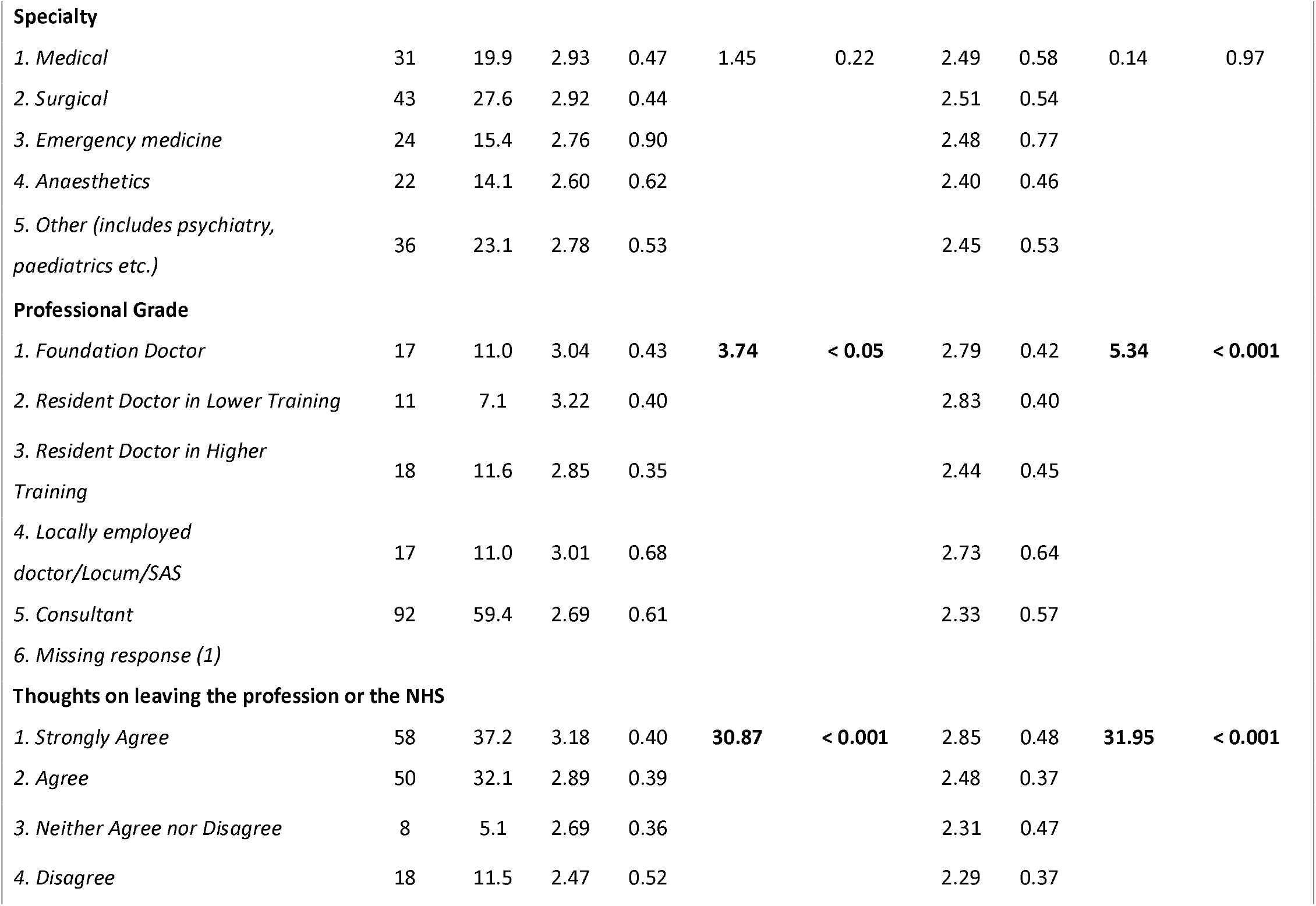
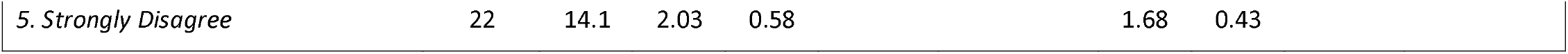
Descriptive statistics, one-way ANOVA, and Tukey post-hoc for socio-demographic variables, and thoughts on leaving the profession or the NHS.

One-way ANOVA identified significant differences in exhaustion scores according to professional grade (*F* = 3.74, *p* = <0.05) and thoughts of leaving the profession or the NHS (*F* = 30.87, *p* < 0.001). Similarly, significant differences were identified in disengagement scores according to professional grade (*F* = 5.34, *p* < 0.001) and thoughts of leaving the profession or the NHS (*F* = 31.95, *p* < 0.001). There were no statistically significant differences in either exhaustion or disengagement scores according to gender, ethnicity, age, or specialty. Following the one-way ANOVA, Tukey’s post-hoc test was undertaken to determine which groups differed significantly within each variable. Resident doctors in lower training were found to have a statistically significantly higher exhaustion and disengagement scores than consultants, while foundation doctors and locally employed doctors/locum/SAS were found to have a statistically significantly higher disengagement score than consultants. Those that answered ‘strongly agree’ to thoughts of leaving the profession or the NHS showed statistically significantly higher mean exhaustion and disengagement scores compared to all other responses.

To determine the extent to which mean exhaustion and disengagement scores were correlated with the socio-demographic variables, and between these scores and on thoughts on leaving the profession or the NHS, correlational analysis was completed using Pearson’s correlation coefficient (Table 3). This showed a statistically significant positive correlation between mean exhaustion and disengagement scores (r = 0.76, *p* < 0.01). Both mean exhaustion and disengagement scores were found to be weakly negatively correlated with both age group (r = -0.20, *p* < 0.01 and r = -0.23, *p* < 0.01) and professional grade (r = -0.26, *p* < 0.01 and r = -0.30, *p* < 0.01), meaning these scores decreased as age or seniority increased. Similarly, both mean exhaustion and disengagement score were found to be strongly positively correlated with thoughts on leaving the profession or the NHS (r = 0.67, *p* < 0.01 and r = -0.65, *p* < 0.01), which indicated as scores for these burnout dimensions increased so did thoughts on leaving the profession or the NHS. Interestingly, a moderate negative correlation was also found between professional grade and thoughts on leaving the profession or the NHS (r = -0.37, *p* < 0.01), indicating individuals who were earlier in their career were more likely to agree with this statement.

**Table 3.**
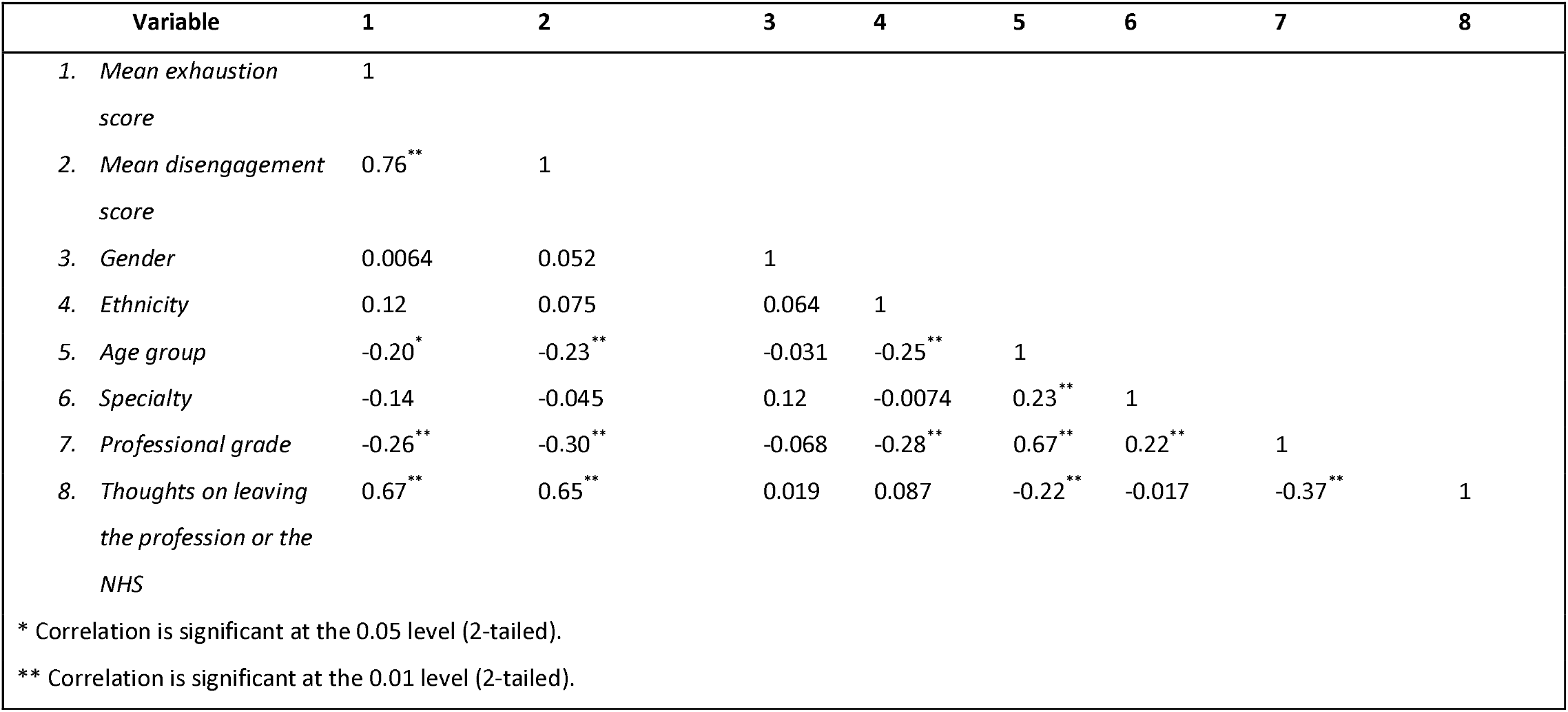
Pearson correlation table of mean exhaustion and disengagement scores, socio-demographic variables, and thoughts on leaving the profession or the NHS.

A linear regression model using the enter method was used to investigate the potentially predictive socio-demographic variables on both exhaustion and disengagement scores (Table 4). Statistical significance was only seen between disengagement score and professional grade, however, the low R^2^ values indicate the model accounts for only 8% and 9% of the variance seen in exhaustion and disengagement score. This model is therefore a poor predictor of these scores.

**Table 4.** Multiple linear regression model between socio-demographic variables and exhaustion and disengagement scores.

| Variable | *B* | *SE* | $\beta$ | *p value* | $R^2$ |
| --- | --- | --- | --- | --- | --- |
| **Exhaustion:** |  |  |  |  | 0.08 |
| *Gender identity* | -0.01 | 0.08 | -0.01 | 0.99 |  |
| *Ethnicity* | 0.07 | 0.10 | 0.06 | 0.50 |  |
| *Age group* | -0.02 | 0.06 | -0.03 | 0.77 |  |
| *Specialty* | -0.04 | 0.03 | -0.09 | 0.27 |  |
| *Professional grade* | -0.08 | 0.05 | -0.20 | 0.07 |  |
| **Disengagement:** |  |  |  |  | 0.09 |
| *Gender identity* | 0.03 | 0.08 | 0.03 | 0.70 |  |
| *Ethnicity* | -0.02 | 0.09 | -0.02 | 0.85 |  |
| *Age group* | -0.03 | 0.06 | -0.05 | 0.64 |  |
| *Specialty* | 0.01 | 0.03 | 0.02 | 0.79 |  |
| *Professional grade* | -0.11 | 0.04 | -0.27 | < 0.05 |  |

As mentioned previously, participants were categorised into burnout groups based on their mean exhaustion and disengagement scores. The largest group of participants (*n* = 111, 71%) were identified as being part of the burnout group (Table 5) and a full breakdown of burnout group by socio-demographic variable and thoughts on leaving the profession or the NHS is shown in Table 6.

**Table 5.**
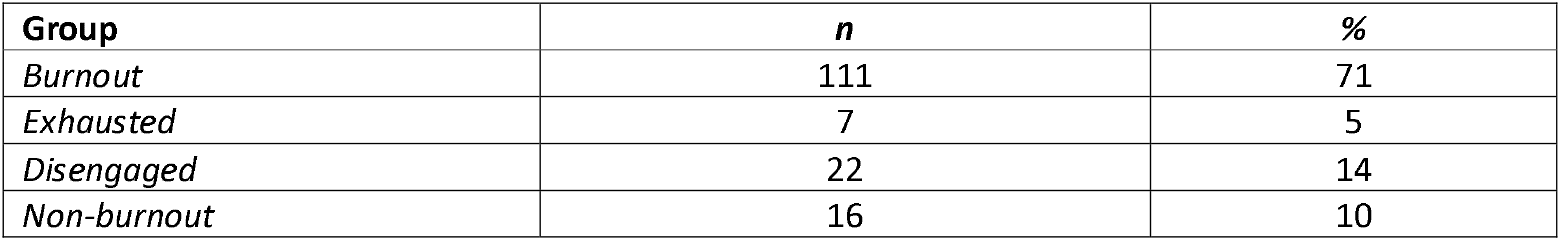
Descriptive statistics of participants by burnout group.

| Group | *n* | % |
| --- | --- | --- |
| *Burnout* | 111 | 71 |
| *Exhausted* | 7 | 5 |
| *Disengaged* | 22 | 14 |
| *Non-burnout* | 16 | 10 |

**Table 6.** Full breakdown of burnout group by socio-demographic variable and thoughts on leaving the profession or the NHS.

| Variable | <u>Burnout group</u> |  | <u>Exhausted group</u> |  | <u>Disengaged group</u> |  | <u>Non-burnout group</u> |  |
| --- | --- | --- | --- | --- | --- | --- | --- | --- |
|  | *n* | % | *n* | % | *n* | % | *n* | % |
| **Gender** |  |  |  |  |  |  |  |  |
| 1. Male | 59 | 53.2 | 15 | 68.2 | 4 | 57.1 | 7 | 46.7 |
| 2. Female | 51 | 45.9 | 7 | 31.8 | 3 | 42.9 | 7 | 46.7 |
| 3. Prefer not to say | 1 | 0.9 | 0 | 0 | 0 | 0 | 1 | 6.7 |
| 4. Missing response (1) |  |  |  |  |  |  |  |  |
| **Ethnicity** |  |  |  |  |  |  |  |  |
| 1. White (includes White British and non-British) | 67 | 60.4 | 14 | 63.6 | 5 | 71.4 | 12 | 75.0 |
| 2. Non-White | 44 | 39.6 | 7 | 31.8 | 2 | 28.6 | 4 | 25.0 |
| 3. Prefer not to say | 0 | 0 | 1 | 4.5 | 0 | 0 | 0 | 0 |
| **Age group** |  |  |  |  |  |  |  |  |
| 1. 20 – 29 | 13 | 11.7 | 0 | 0.0 | 0 | 0.0 | 2 | 12.5 |
| 2. 30 - 39 | 41 | 36.9 | 7 | 31.8 | 1 | 14.3 | 4 | 25.0 |
| 3. 40 - 49 | 36 | 32.4 | 6 | 27.3 | 2 | 28.6 | 6 | 37.5 |
| 4. 50 - 59 | 18 | 16.2 | 7 | 31.8 | 2 | 28.6 | 4 | 25.0 |
| 5. 60 and over | 3 | 2.7 | 1 | 4.5 | 2 | 28.6 | 0 | 0 |
| 6. Prefer not to say | 0 | 0 | 1 | 4.5 | 0 | 0 | 0 | 0 |
| **Specialty** |  |  |  |  |  |  |  |  |
| 1. Medical | 23 | 20.7 | 5 | 22.7 | 0 | 0.0 | 3 | 18.8 |
| *2. Surgical* | 32 | 28.8 | 8 | 36.4 | 1 | 14.3 | 2 | 12.5 |
| *3. Emergency medicine* | 18 | 16.2 | 1 | 4.5 | 0 | 0 | 5 | 31.3 |
| *4. Anaesthetics* | 12 | 10.8 | 4 | 18.2 | 3 | 42.9 | 3 | 18.8 |
| *5. Other (includes psychiatry, paediatrics etc.)* | 26 | 23.4 | 4 | 18.2 | 3 | 42.9 | 3 | 18.8 |
| **Professional Grade** |  |  |  |  |  |  |  |  |
| *1. Foundation Doctor* | 15 | 13.6 | 0 | 0 | 0 | 0 | 2 | 12.5 |
| *2. Resident Doctor in Lower Training* | 10 | 9.1 | 1 | 4.5 | 0 | 0 | 0 | 0 |
| *3. Resident Doctor in Higher Training* | 14 | 12.7 | 4 | 18.2 | 0 | 0 | 0 | 0 |
| *4. Locally employed doctor/Locum/SAS* | 14 | 12.7 | 1 | 4.5 | 1 | 14.3 | 1 | 6.3 |
| *5. Consultant* | 57 | 51.8 | 16 | 72.7 | 6 | 85.7 | 13 | 81.3 |
| *6. Missing response (1)* |  |  |  |  |  |  |  |  |
| **Thoughts on leaving the profession or the NHS** |  |  |  |  |  |  |  |  |
| *1. Strongly Agree* | 53 | 47.7 | 4 | 18.2 | 0 | 0 | 1 | 6.3 |
| *2. Agree* | 40 | 36.0 | 7 | 31.8 | 2 | 28.6 | 1 | 6.3 |
| *3. Neither Agree nor Disagree* | 4 | 3.6 | 3 | 13.6 | 1 | 14.3 | 0 | 0 |
| *4. Disagree* | 11 | 9.9 | 3 | 13.6 | 3 | 42.9 | 1 | 6.3 |
| *5. Strongly Disagree* | 3 | 2.7 | 5 | 22.7 | 1 | 14.3 | 13 | 81.3 |

To account for the limited data seen in the contingency tables, the association between burnout groups and each socio-demographic variable, as well as the association between burnout groups and thoughts of leaving the profession or the NHS, was analysed using Fisher-Freeman-Halton tests (Table 7).

**Table 7:** Fisher-Freeman-Halton test between socio-demographic variables and burnout groups, and thoughts of leaving the profession or the NHS and burnout groups.

| Variable | Value | *p value* |
| --- | --- | --- |
| *Gender identity* | 5.91 | 0.44 |
| *Age group* | 20.48 | 0.11 |
| *Ethnicity* | 7.60 | 0.37 |
| *Professional grade* | 12.39 | 0.29 |
| *Specialty* | 14.51 | 0.20 |
| *Thoughts of leaving the profession or the NHS* | 68.42 | < 0.001 |

While this showed no statistically significant association between burnout group and socio-demographic variable, there was a statistically significant association between burnout group and thoughts of leaving the profession or the NHS (*p* < 0.001).

Similarly, due to the limited data seen in the contingency tables, the ‘Exhausted’, ‘Disengaged’, and ‘Non-burnout’ groups were grouped into ‘Not burnout’ and a binary linear regression model using the enter method was used to investigate the potentially predictive socio-demographic variables on burnout (Table 8).

**Table 8:**
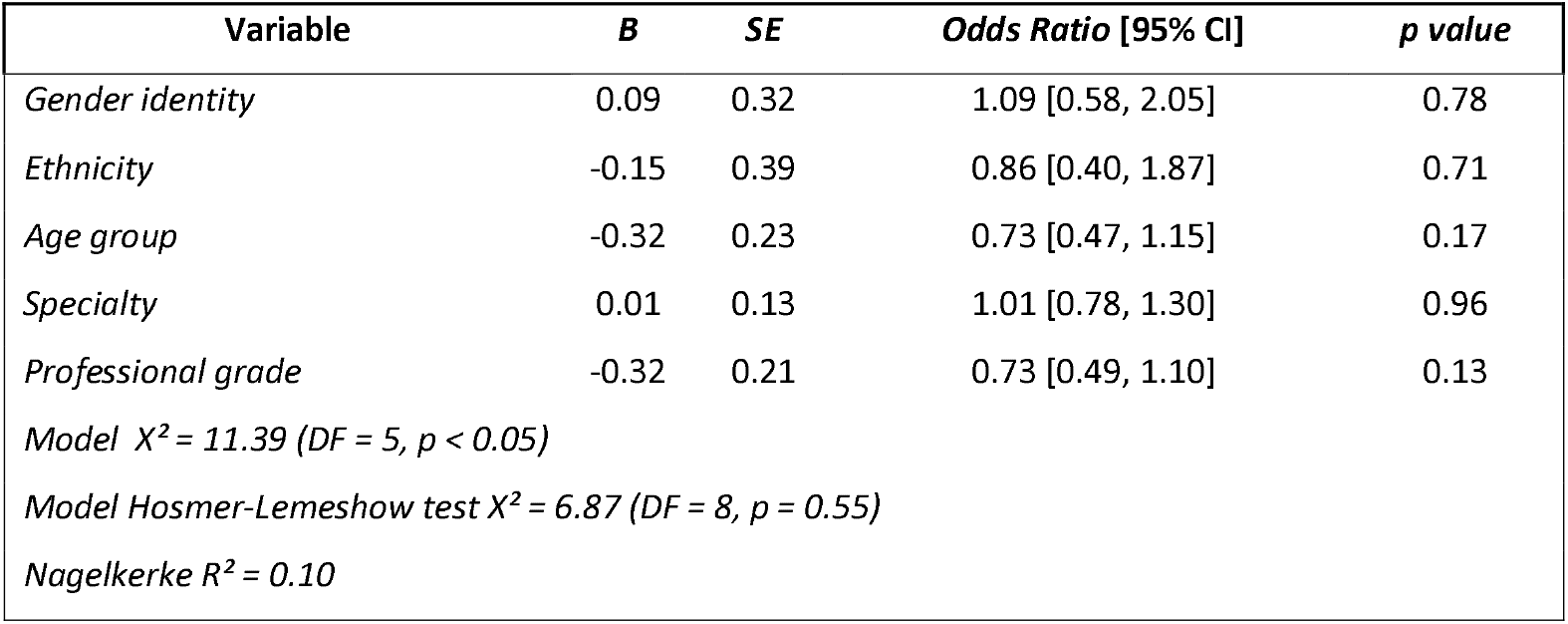
Binary linear regression results to determine the socio-demographic variables which predict burnout.

| Variable | *B* | *SE* | *Odds Ratio* [95% CI] | *p value* |
| --- | --- | --- | --- | --- |
| *Gender identity* | 0.09 | 0.32 | 1.09 [0.58, 2.05] | 0.78 |
| *Ethnicity* | -0.15 | 0.39 | 0.86 [0.40, 1.87] | 0.71 |
| *Age group* | -0.32 | 0.23 | 0.73 [0.47, 1.15] | 0.17 |
| *Specialty* | 0.01 | 0.13 | 1.01 [0.78, 1.30] | 0.96 |
| *Professional grade* | -0.32 | 0.21 | 0.73 [0.49, 1.10] | 0.13 |
| *Model $X^2 = 11.39$ (DF = 5, $p < 0.05$)* |  |  |  |  |
| *Model Hosmer-Lemeshow test $X^2 = 6.87$ (DF = 8, $p = 0.55$)* |  |  |  |  |
| *Nagelkerke $R^2 = 0.10$* |  |  |  |  |

The overall model was statistically significant when compared to the null model (*X*^2^ = 11.39, *p* < 0.05), indicating the socio-demographic variables, when modelled together, are significant predictors of burnout. The Hosmer-Lemeshow test (*X*^2^ = 6.87, *p* = 0.55) indicates there is an acceptable fit of the model to the observed data. However, these variables only explained 10% of the variance seen in burnout (Nagelkerke *R*^*2*^). This suggests that the model’s ability to predict burnout based on these variables is limited, and other variables which have not been identified may play a role in burnout.

## Discussion

This study sought investigate burnout among doctors working at a major West Midlands trauma centre using the widely validated OLBI as the primary evaluation tool. This study revealed a strong association between exhaustion and disengagement dimensions of burnout and both professional grade and thoughts of leaving the profession or the NHS (Table 2). Post-hoc analysis highlighted that scores for both these dimensions were significantly lower for consultants compared to doctors who were in the earlier stages of their medical career. This was supported by correlational analysis which showed a weak negative correlation between these burnout dimensions and professional grade, indicating burnout decreased as an individual gained more seniority, and this is consistent with previous work (Balendran et al., 2021).

While the responding sample of 156 doctors found 71% were identified as meeting the criteria for burnout, no individual socio-demographic variable was found to be statistically associated with burnout group (Table 7) or be a significant predictor of burnout (Table 8). However, the binary linear regression model (Table 8) determined that while socio-demographic variables, collectively, are predictors of burnout, only 10% of the variation in burnout is accounted for by these variables. The statistically significant association found between burnout group and thoughts on leaving the profession or the NHS (Table 6) may indicate that factors not captured in this study such as those external to a doctor’s locus of control, like staffing levels, play a key role in burnout rather than socio-demographic variables, and this is consistent with previous studies that have looked at burnout among doctors (Nwosu et al., 2020; Nayer et al., 2024; Donald and Lindsay, 2024). Employers, therefore, need to recognise this and implement organisation-wide strategies that can help minimise burnout among doctors, particularly those who are in the earlier stages of their career. Future studies should seek to investigate the impact of organisation culture and work environment on burnout and the effectiveness of targeted interventions, which could include improving on work conditions by providing adequate rest facilities, free staff parking, or by increasing staff retention via more comprehensive renumeration packages, in order to reduce burnout. Additionally, it is important that employers are aware that burnout among doctors does not just affect the individual doctor, but also has wider implications. A systematic review by Wilkinson et al. (2017) identified a negative association between burnout and empathy towards patients, and another by Panagioti et al. (2018) demonstrated burnout was associated with an increased risk of patient safety incident events.

There were several limitations in this study. Firstly, the number of respondents to the survey was relatively low which meant the generalisability of the findings were limited, particularly as the individual burnout groups had to be recategorised in order to determine if there were any potential predictive variables on burnout, and response rates could not be calculated due to the fluctuating number of doctors at the hospital. Secondly, due to the nature of how the survey was distributed to participants, selection bias may have been introduced as this study could preferentially attract those who have a particular interest in burnout or who have first or second-hand experience of it, and this may have resulted in a disproportionally high number of consultants who participated in this study. Finally, the cross-sectional nature of the study may have led to the introduction of response bias as participants may not be able to accurately recall their previous experiences with burnout. This could have been remedied by utilising both quantitative and qualitative data or by holding interviews with participants in order to obtain a deeper understanding of their experience with burnout, or by conducting a longitudinal study to better understand how the exhaustion and disengagement dimensions, and therefore burnout, evolve over time.

## Conclusion

At University Hospital Coventry, burnout is a persistent and prevalent issue that affects doctors across all societal and demographic groups. While 71% of doctors who took part in this study were found to be experiencing burnout, as reflected by high scores in both exhaustion and disengagement, doctors who were earlier in their careers were found to have statistically significant higher scores in both these burnout dimensions compared to more senior doctors. Furthermore, those experiencing burnout had a statistically strong association with thoughts of leaving the profession or the NHS. It is crucial, therefore, that organisations must focus on implementing targeted interventions that minimise burnout in order to minimise the workforce crisis currently plaguing the UK healthcare system. Additionally, this study shows that there is value in collecting socio-demographic data such as professional grade when assessing burnout and recommends that the GMC adopts the use of the OLBI to better evaluate and extract insights in burnout that can be used for workforce planning.

## Data Availability

All data in the present study are available upon reasonable request to the authors.

## Appendix A

Oldenburg Burnout Inventory. The (R) denotes questions which are reversed and means that a higher score indicates a higher level of burnout.

**Exhausted:**

*There are days when I feel tired before I arrive at work (R)*

*After work, I tend to need more time than in the past in order to relax and feel better (R)*

*I can tolerate the pressure of my work very well*

*During my work, I often feel emotionally drained (R)*

*After working, I have enough energy for my leisure activities*

*After my work, I usually feel worn out and weary (R)*

*Usually, I can manage the amount of my work well*

*When I work, I usually feel energized*

**Disengaged:**

*I always find new and interesting aspects in my work*

*It happens more and more often that I talk about my work in a negative way (R)*

*Lately, I tend to think less at work and do my job almost mechanically (R)*

*I find my work to be a positive challenge*

*Over time, one can become disconnected from this type of work (R)*

*Sometimes I feel sickened by my work tasks (R)*

*This is the only type of work that I can imagine myself doing I feel more and more engaged in my work*

## References

Aiello, E.N., Fiabane, E., Margheritti, S., Magnone, S., Bolognini, N., Miglioretti, M. & Giorgi, I. (2022b) Psychometric properties of the Copenhagen Burnout Inventory (CBI) in Italian Physicians. Med Lav, 113(4):e2022037.

Balendran, B., Bath, MF, Awopetu, A. & Kreckler, SM (2021) Burnout within UK surgical specialties: a systematic review. Ann R Coll Surg Engl, 103(7):464–470.

Cordero-Franco, HF, Salinas-Martínez, AM, Chávez-Barrón, KA, Espinoza-Torres, FG, Guzmán-de la Garza, FJ & Moreno-Treviño, CA (2022) Validation of the Spanish Version of the Copenhagen Burnout Inventory in Mexican Medical Residents. Arch Med Res, 53(6):617–624.

Demerouti, E. and Bakker, A. B. (2008) ‘The Oldenburg Burnout Inventory: A good alternative to measure burnout and engagement’, Handbook of Stress and Burnout in Health Care, 65–78

Donald, N. and Lindsay, T. (2024) Surgical trainee experiences from 2013 to 2023 within the United Kingdom as reported by the General Medical Council National Training Survey. Surgeon, 22(2):74–79.

Faul, F., Erdfelder, E., Buchner, A. & Lang, AG (2009) Statistical power analyzes using G*Power 3.1: tests for correlation and regression analyses. Behav Res Methods, 41 (4): 1149–1160.

General Medical Council (2024a) National training survey 2024 report. Available at: https://www.gmc-uk.org/-/media/documents/national-training-survey-summary-report-2024_pdf-107834344.pdf (accessed: 26 November 2024)

General Medical Council (2024b) The state of medical education and practice in the UK. Workplace experiences 2024. Available at https://www.gmc-uk.org/-/media/documents/somep-workplace-report-2024-full-report_pdf-107930713.pdf (accessed: 26 November 2024)

Halbesleben, J. R. B. and Demerouti, E. (2005) ‘The construct validity of an alternative measure of burnout: Investigating the English translation of the Oldenburg Burnout Inventory’, Work & Stress, 19(3), pp. 208–220. doi: 10.1080/02678370500340728.

IBM Corp. (2023). IBM SPSS Statistics for Windows, Version 29.0. Armonk, NY: IBM Corp. https://www.ibm.com/products/spss-statistics

Khan, A. and Yusoff, R. (2016) ‘Psychometric Testing of Oldenburg Burnout Inventory among Academic Staff in Pakistan’, International Review of Management and Marketing, 6, pp. 683–687

Kristensen, T. S., Borritz, M., Villadsen, E. and Christensen, K. B. (2005) ‘The Copenhagen Burnout Inventory: A new tool for the assessment of burnout’, Work & Stress, 19(3), pp. 192–207. doi: 10.1080/02678370500297720.

Nayar, S.K., Acquaah, F., Kayani, B. & Vemulapalli, K. (2024) Burnout in trauma and orthopedics: a cross-sectional study of surgeons from across the United Kingdom. Ann R Coll Surg Engl, 106(2):131–139.

Nwosu, ADG, Ossai, EN, Mba, UC, Anikwe, I., Ewah, R., Obande, B. & Achor, JU (2020) Physician burnout in Nigeria: a multicentre, cross-sectional study. BMC Health Serv Res, 20(1):863.

Panagioti, M., Geraghty, K., Johnson, J., Zhou, A., Panagopoulou, E., Chew-Graham, C., Peters, D., Hodkinson, A., Riley, R. & Esmail, A. (2018) Association Between Physician Burnout and Patient Safety, Professionalism, and Patient Satisfaction: A Systematic Review and Meta-analysis. JAMA Intern Med, 178(10):1317–1331.

Peterson, U., Demerouti, E., Bergström, G., Samuelsson, M., Asberg, M. & Nygren, A. (2008) Burnout and physical and mental health among Swedish healthcare workers. J Adv Nurs, 62(1):84–95.

Podsakoff, PM, MacKenzie, SB, Lee, J. & Podsakoff, NP (2003) Common method biases in behavioral research: a critical review of the literature and recommended remedies. J Appl Psychol, 88(5):879–903.

Schaufeli, W.B., Bakker, AB, Hoogduin, K., Schaap, C. & Kladler, A. (2001) on the clinical validity of the maslach burnout inventory and the burnout measure. Psychol Health, 16 (5): 565–582.

Tipa, R.O., Tudose, C. & Pucarea, V.L. (2019) Measuring Burnout Among Psychiatric Residents Using the Oldenburg Burnout Inventory (OLBI) Instrument. J Med Life, 12(4):354–360.

Wilkinson, H., Whittington, R., Perry, L. & Eames, C. (2017) Examining the relationship between burnout and empathy in healthcare professionals: A systematic review. Burn Res, 6 18–29.

